# An observational study of uptake and adoption of the NHS App in England

**DOI:** 10.1101/2022.03.16.22272200

**Authors:** Salina Tewolde, Céire Costelloe, John PowelI, Chrysanthi Papoutsi, Claire Reidy, Bernard Gudgin, Craig Shenton, Felix Greaves

## Abstract

**Objectives:** This study aimed to evaluate patterns of uptake and adoption of the NHS App. Data metrics from the NHS App were used to assess acceptability by looking at total app downloads, registrations, appointment bookings, GP health records viewed, and prescriptions ordered. The impact of the UK COVID-19 lockdown and introduction of the *COVID Pass* were also explored to assess App usage and uptake.

**Methods:** Descriptive statistics and an interrupted time series analysis were used to look at monthly NHS App metrics at a GP practice level from January 2019-May 2021 in the population of England. Interrupted time series models were used to identify changes in level and trend among App usage and the different functionalities before and after the first COVID-19 lockdown. The *Strengthening the Reporting of Observational Studies in Epidemiology (STROBE)* guidelines were used for reporting and analysis.

**Results:** Between January 2019 and May 2021, there were a total of 8,524,882 NHS App downloads and 4,449,869 registrations. There was a 4-fold increase in app downloads from April 2021 (650,558 downloads) to May 2021 (2,668,535 downloads) when the COVID Pass feature was introduced. Areas with the highest number of App registrations proportional to the GP patient population occurred in Hampshire, Southampton and Isle of Wight CCG, and the lowest in Blackburn with Darwen CCG. After the announcement of the first lockdown (March 2020), a positive and significant trend in the number of login sessions was observed at 602,124 (p=0.004)** logins a month. National NHS App appointment bookings ranged from 298 to 42,664 bookings per month during the study period. The number of GP health records viewed increased by an average of 371,656 (p=0.001)** views per month and the number of prescriptions ordered increased by an average of 19934 (p<0.001)*** prescriptions per month following the first lockdown.

**Conclusion:** This analysis has shown that uptake and adoption of the NHS App was positive post lockdown, and increased significantly due to the COVID Pass feature being introduced, but further research is needed to measure the extent to which it improves patient experience and influences health service access and care outcomes.

## Introduction

The increasing adoption of electronic medical records (EMRs) by hospitals and GPs have given patients digital access to their personal health information through the utilization of patient portals. Patient portals provide the potential for improved patient-provider communication via electronic messaging, direct appointment booking, prescription ordering, and provide users a platform to be more informed about their health.^1^ Some preliminary evidence has shown that patient portals may improve health outcomes and reduce health inequities, but the direct benefits are still unclear.^9,10,11,12^ There are also issues around trust, security, communication and interoperability with these new technologies.^19^ Work by the OpenNotes team in the US suggests that offering patients access to doctors’ notes is acceptable, improves communication and patients’ confidence in managing their care, but also finds many clinicians are still resistant to the idea.^20,21,22^ Recent reviews on patient portals show their widespread use in other countries and that patients’ interest and ability to use them is influenced by age, ethnicity, education, health literacy, and health status.^23^ One example of an established patient portal in the United States, is the Kaiser Permanente Northern California (KPNC)’s inpatient and ambulatory care electronic health record system. KPNC’s 3.4 million patient portal users use the platform to exchange messages with their provider, create appointments, refill prescriptions, and view their medical records.^13^ Other national patient portals have been established in Denmark, Estonia and Australia, where all citizens currently have access to their personal health data, but there is still uncertainty surrounding the impact on these portals and care outcomes and patient experience.^24^

### The NHS App

In January 2019, the National Health Service (NHS) in England introduced a new app for patients, called “the NHS App”. The enabled functions of the App for general use at launch were:

1. Check symptoms using NHS 111 online and the health A-Z on the NHS website

2. View their GP medical record

3. Book and manage appointments at their GP practice

4. Order repeat prescriptions

5. Register as an organ donor

6. Set data sharing preferences for the national data: choose if data is shared for planning and research

In light of the novel coronavirus (COVID-19) pandemic, a new feature called the NHS “COVID Pass” was announced on April 28, 2021 and was then launched on May 17, 2021 under the name “NHS COVID Pass”. The COVID Pass allowed users to prove their vaccination status in England, and received great interest in the UK media during the launch.^25^ Users were also able to use the “Check your COVID-19 vaccine record” feature on the App which allowed patients to see which vaccines they had received historically.

With over 3,000,000 mobile health applications currently on the market, a gap remains in identifying validated digital health solutions and their impact on public health. NHS England’s goals for the App are to 1) improve access to primary care services, 2) improve patient experience, 3) save time in GP practices and 4) promote self-care. Our research aims to evaluate the effectiveness of the NHS App in meeting its goals by looking at early patterns of uptake and adoption. We will use observational data from the NHS App to assess use and acceptability by looking at total app downloads and registrations, total number of appointments bookings, total number of GP health records viewed, and the number of prescriptions ordered. We will also look at the change in frequency amongst these functionalities due to the impact of COVID-19 and the introduction of the COVID Pass.

## Methods

### Data

Data from the NHS App were analyzed using data metrics from the NHS App Dashboard.^18^ The NHS App Dashboard was jointly developed by NHS England and Improvement team and NHS Digital under England’s National Health Care system.^15^

In this study, aggregate counts of NHS App activity were analyzed using data sourced from the NHS App Dashboard.^18^ The NHS App Dashboard was jointly developed by NHS England and Improvement team and NHS Digital.^15^ Metrics currently available through the dashboard include aggregate counts of the number of registrations, downloads, appointment bookings and cancellations, GP health records viewed, prescription requests, visits to NHS 111 online, organ donation registrations and withdrawals, visits to the health A-Z page, and visits to the national data opt-out page. These metrics can be viewed on a weekly or monthly basis and can be broken down geographically, by NHS Regions, Sustainability and Transformation Partnerships (STP) (planning framework for NHS services), Clinical Commissioning Groups (CCG) (set up by the Health and Social Care Act 2012 that deliver NHS services in England), and by GP practice.^6^

#### NHS App data processing

Anonymous analytics logs from the NHS App iPhone and Android applications service are collected and stored on a secure cloud service by NHS Digital. The NHS App data team at NHS Digital then generates an aggregate GP practice level count of app activity on a weekly/monthly basis.^18^ The aggregate data are simply counts of the number of times each app feature was used in a given geography. These aggregate data are then used to develop the NHS App dashboard which is deployed on the NHS England Applications platform. At no point is patient level data made available or used by analysts at NHSX or Imperial College London. The research team did not have access to any personal or personally identifiable information. All NHS App metrics and descriptions are included in the Appendix.

### Analysis

#### Descriptive statistics

The study period for this analysis was from January 2019 to May 2021. Descriptive statistics were used to summarize monthly NHS App metrics at a GP practice level for the entire country. An absolute monthly count was presented from January 2019 to May 2021 of total App downloads, registrations, login sessions, appointments booked, GP health records viewed, and prescriptions ordered. New App downloads and registrations were also presented during the same time frame. A heat map extracted from the NHS App Dashboard displays the total percentage of the GP patient population aged 13+ registered for the App across the whole country. Lowest and highest App registrations by CCG were also presented for the population of England.

#### Time Series Analysis

An ecological time series and an interrupted time series (ITS) analysis was used to analyze the impact of the first UK national lockdown due to COVID-19 that occurred on March 26, 2020. The time series analysis was used to evaluate changes in uptake and the longitudinal impact of the pandemic on different functionalities of the App.^7^ An interruption was added to the time series on April 1, 2020, to estimate changes in uptake before and after announcement of the first lockdown. The ITS explored changes in level and trend for national App logins, appointment bookings, GP health records viewed, and prescriptions ordered between January 2019 and May 2021. For each App metric, a linear regression model with a time series specification was used, where pre-lockdown was denoted by “0” and the post-lockdown period was denoted by “1”.

The ITS model was then used to assess if there was a change in App usage immediately after the first lockdown (the level) and if a change in trend occurred over the whole study period. Absolute and relative changes of the post intervention trend were also estimated, had the first lockdown not taken place. The absolute change in trend was calculated by taking the difference between the predicted pre-lockdown trend of the outcome and the post-lockdown trend at the end of the study period. The relative change was calculated as the absolute change as a relative proportion. To analyze App usage and uptake before the NHS COVID Pass was introduced, a separate ITS analysis was conducted excluding the month of May 2021. Autocorrelation was assessed by analyzing the residuals of the ITS models.^8^ The autocorrelation function plots (ACF) and partial autocorrelation function (PACF) plots of the residuals are included in the Appendix.

The time series model for App metric Y at time *t* had the form:

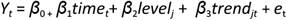

Where for each app metric *Y*,

***β***_***0***_ estimates the baseline count of app metrics at time 0 (January 2019)

***β***_***1***_ estimates the baseline trend before the first lockdown was implemented

***β***_*2*_ estimates the change in level in app metrics immediately after the first lockdown

***β***_*3*_ estimates the change in trend after the first lockdown was implemented (change in slope)

***t*** represents time in months from the start to the end of the study period (Jan 2019-May 2021)

***e*** represents the residual for the model

All analyses were performed using R Studio version 1.4.1717 (RStudio, Northern Avenue, MA, US). The regression coefficients obtained from the ITS were considered significant if p<0.005. The Strengthening the Reporting of Observational Studies in Epidemiology (STROBE) guidelines were used for the reporting and analysis of this study.^5^

## Results

### NHS App Registrations and Download

There were a total of 8,524,882 NHS App downloads and 4,449,869 registrations from January 2019 to May 2021. At the start of the study period, there were 31,633 downloads and 1,279 registrations. At the onset of the first UK lockdown in March 2020, there was a spike of 532,275 App downloads. The highest number of downloads during the study period occurred after the announcement and launch of the COVID Pass in May 2021, with a total of 2,668,535 downloads. During this month, 2,099,234 users registered for the App. At the end of May 2021, there were 51,956,423 GP registered patients in England. Of these registered patients, 8.56% of the population aged 13+ were registered for the NHS App. **Figure 1** illustrates the monthly App downloads and registrations from January 2019-May 2021.

**Figure 1:**
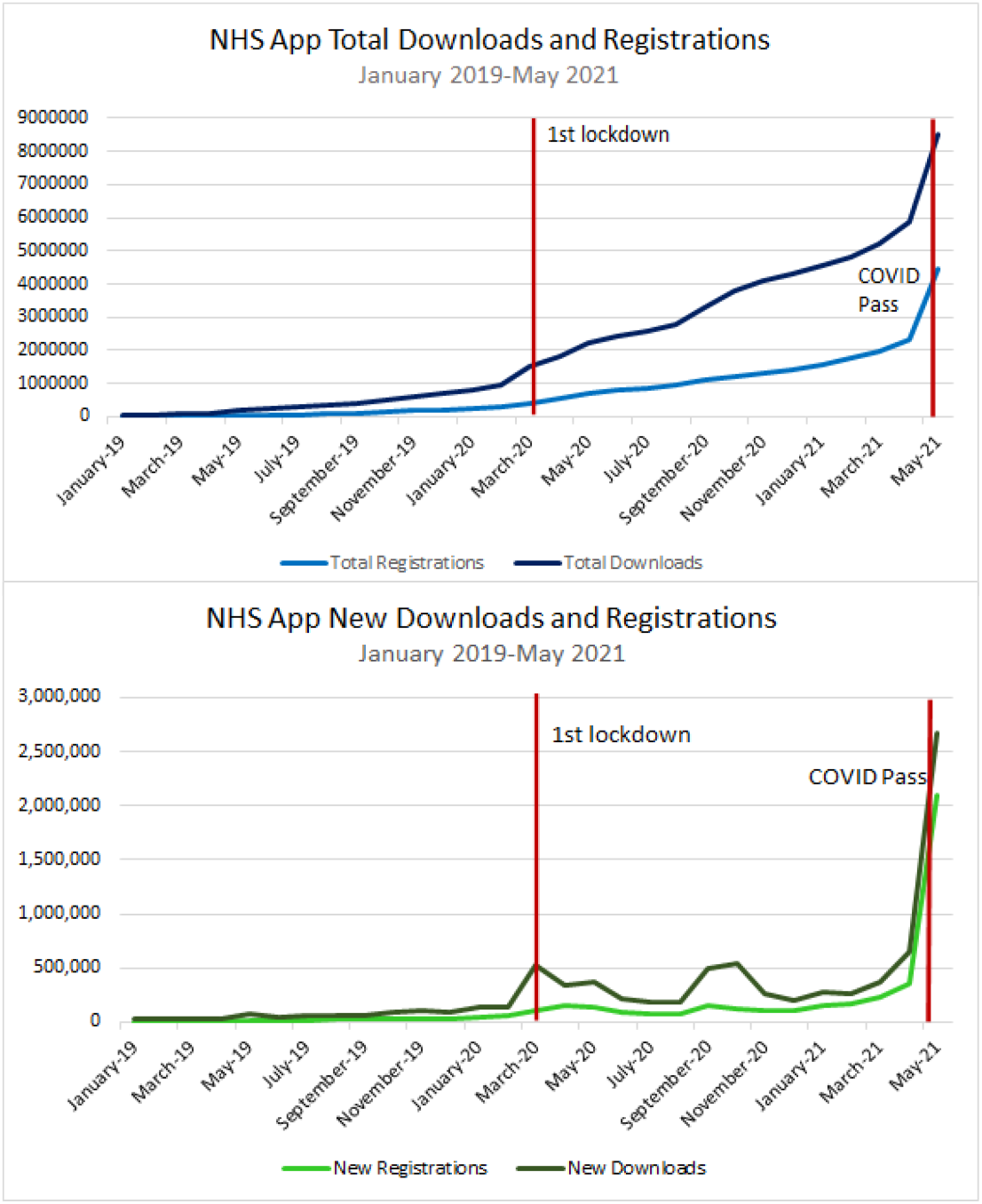
Monthly cumulative and new NHS app registrations and downloads.

Figure 2 illustrates a heat map analysis of the percentage of the GP patient population registered for the NHS app by CCG across England. Registrations ranged from 5.67%-17.08% across the whole country. CCGs with the highest proportion of App registrations occurred in East Riding of Yorkshire, Vale of York, and Hampshire, Southampton and Isle of Wight (15.25%-17.08%). The lowest occurred in Blackburn with Darwen, Morecambe Bay, and Blackpool (5.77%-6.74%).

**Figure 2:**
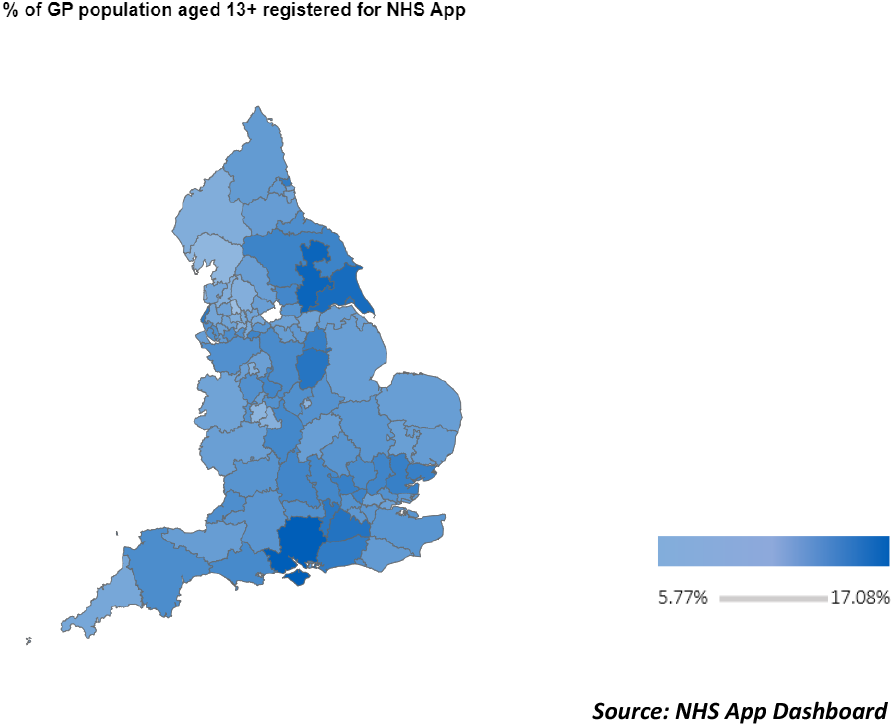
Map of GP population registered for NHS App in England.

### NHS App Metrics

#### Login Sessions

National NHS App login sessions per month ranged from 5,157 to 16,730,430 from January 2019 to May 2021. There was a steady increase of login sessions from January 2019 to April 2021, with the highest number of logins occurring after introduction of the COVID Pass in May 2021 (16,730,430 login sessions in that month) (**Figure 3**). The ITS showed that before implementation of the first lockdown, the number of NHS App login sessions was increasing over time at a rate of 52,093 logins a month, and decreased immediately after implementation of the first lockdown. However, only the change in trend over time was found to be positive and significant, indicating an average slope change of 602,124 (p=0.004) logins a month. 12 months after the first lockdown, the average number of NHS App logins was 5,663,224 more than would have been expected if lockdown did not occur. This represented a 440% increase. (**Table 1**).

**Figure 3:**
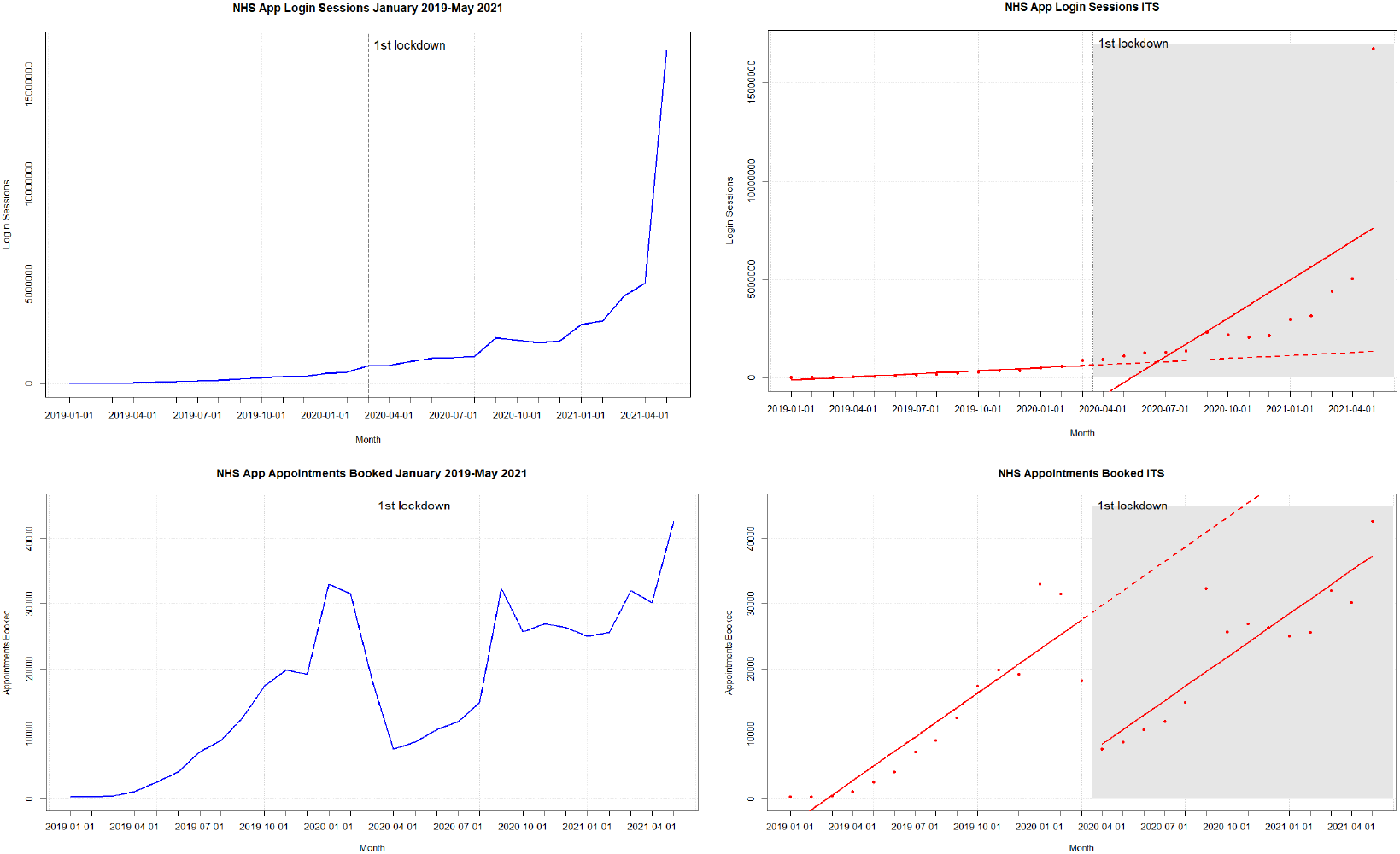
Login Sessions & Appointments Booked time series and Interrupted Times Series (January 2019-May 2021)

**Table 1.** Highest & Lowest NHS App Registrations by CCG in England.

| UK CCGs | % of GP pop aged 13+ registered for NHS App | NHS App Registrations | GP Registered pop aged 13+ years | Total GP list size (from Patient Online) |
| --- | --- | --- | --- | --- |
| Hampshire, Southampton And Isle Of Wight | 17.08% | 248,286 | 1,453,884 | 1,622,560 |
| Vale of York | 16.10% | 52,037 | 323,152 | 364,598 |
| East Riding of Yorkshire | 15.25% | 41,003 | 268,856 | 307,069 |
| Surrey Heartlands | 14.89% | 140,634 | 928,214 | 1,103,431 |
| Hull | 14.27% | 36,746 | 257,591 | 302,845 |
| Frimley | 14.23% | 96,272 | 653,311 | 801,839 |
| Nottingham And Nottinghamshire | 14.10% | 136,688 | 954,206 | 1,106,748 |
| Southport and Formby | 13.97% | 15,418 | 110,395 | 125,591 |
| West Sussex | 13.86% | 112,425 | 810,936 | 904,503 |
| North Tyneside | 13.73% | 27,125 | 197,503 | 222,181 |
| North Cumbria | 7.73% | 22,823 | 295,131 | 326,354 |
| East Lancashire | 7.68% | 25,239 | 328,729 | 384,324 |
| Stoke On Trent | 7.48% | 18,854 | 252,098 | 296,246 |
| Salford | 7.48% | 19,357 | 96,652 | 280,465 |
| Birmingham And Solihull | 7.44% | 83,142 | 1,089,775 | 1,335,087 |
| Leicester City | 6.97% | 24,636 | 353,702 | 423,338 |
| Black Country And West Birmingham | 6.78% | 85,796 | 1,252,576 | 1,486,819 |
| Blackpool | 6.74% | 10,162 | 150,878 | 174,763 |
| Morecambe Bay | 6.69% | 21,413 | 319,958 | 349,652 |
| Blackburn with Darwen | 5.77% | 8,570 | 148,443 | 178,504 |

#### Appointments Booked

Appointments booked using the NHS App ranged from 298 to 42,664 per month during the study period. There was a surge of appointment bookings in January 2020 (33,003 appointments booked), September 2020 (32,335 appointments booked), and the highest in May 2021 (42,644 appointments booked). There was a significant drop in appointment bookings after the first lockdown was announced in April 2020 (7,674 appointments booked). The ITS showed that before the first lockdown, there was significant evidence of an increase in the average number of appointment bookings 2,247 (p<0.001), followed by an immediate decrease in bookings after the lockdown was announced −21,315 (p<0.001). 12 months after the first lockdown, the average number of appointments booked using the NHS App was 21,601 fewer than would have been expected if lockdown did not occur. This represented an overall 38% decrease in appointment bookings. Time series plots and the ITS analysis for NHS App login sessions and appointment bookings are depicted in **Table 1** and **Figure 3**.

#### GP Health Records Viewed

GP health records viewed in the NHS App ranged from 2,212 to 9,324,546 per month from January 2019 to May 2021. There was a steady increase in records viewed up until April 2021, where there was a threefold increase from the months of April 2021 (3,309,586 records viewed) and May 2021 (9324546 records viewed) (**Figure 4**). The ITS showed that the average number of health records viewed right after lockdown declined −1441297 (p=0.11), although not found to be significant. The sustained effect over time was significant (p=0.001) indicating that the number of records viewed increased on average by 371,656 views per month post-lockdown. (**Table 2**) 12 months after the first lockdown, the average number of GP health records viewed was 3,390,234 more than would have been expected if lockdown did not occur. This represented a 548% increase.

**Figure 4:**
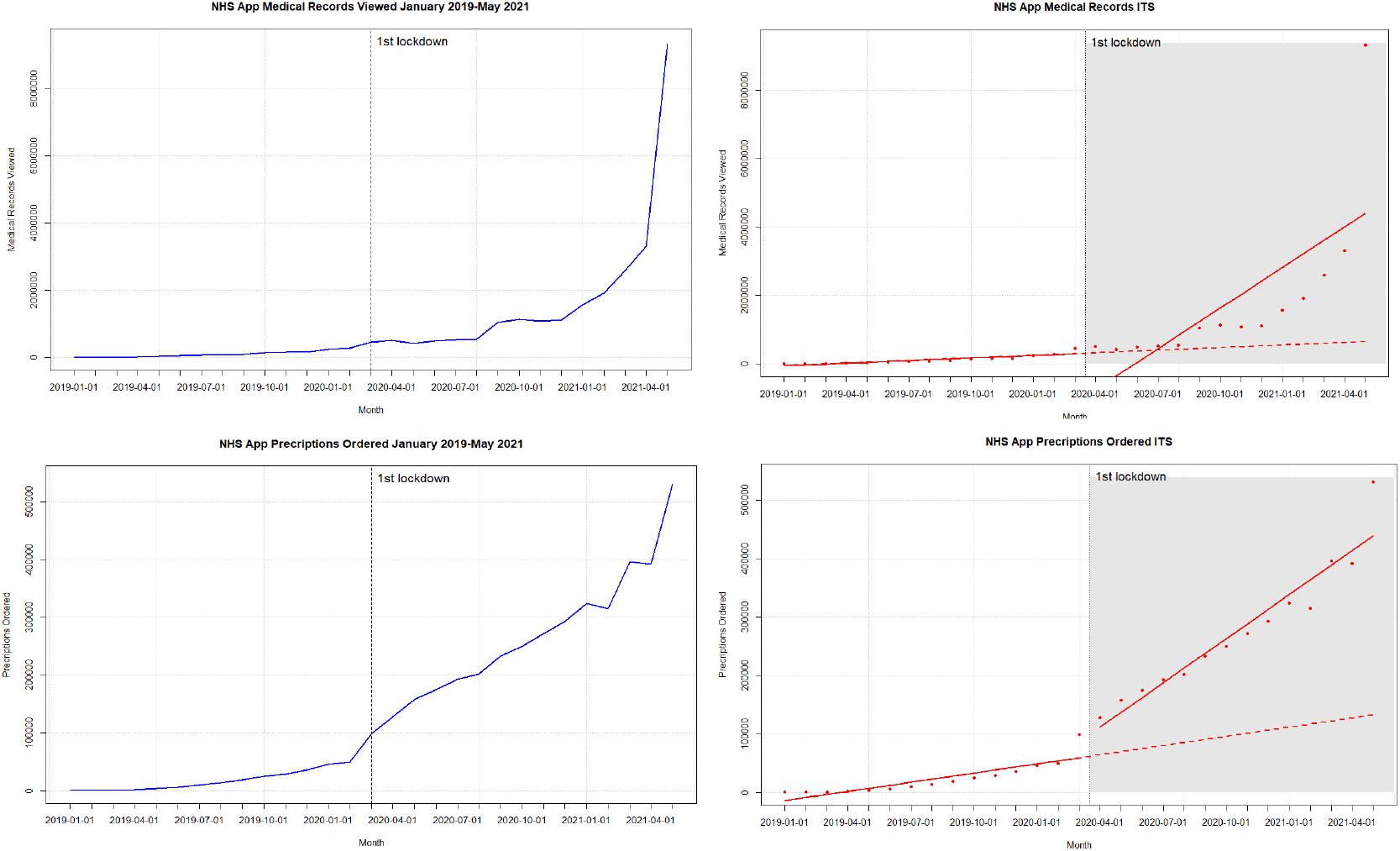
GP Health Records Viewed & Prescriptions Ordered time series and Interrupted Times Series (January 2019-May 2021)

**Table 2.** Login Sessions & Appointments Booked Interrupted Times Series Output.

| NHS App Metric | Regression Intercept<br>(P-value) | Pre-Intervention Trend<br>(P-value) | Immediate Change in Trend after COVID-19 Lockdown<br>(P-value) | Change in Trend over Study Period<br>(P-value) | 1 year after first lockdown |  |
| --- | --- | --- | --- | --- | --- | --- |
|  |  |  |  |  | Absolute Change (count) | Relative Change (percent) |
| Login Sessions<sup>1</sup> | -175074<br>(P=0.88) | 52093<br>(P=0.68) | -2164386<br>(P=0.19) | 602124**<br>(P=0.004) | 5663224 | 441% |
| Login Sessions<sup>2</sup> | -175074<br>(P=0.36) | 52093<br>(P=0.02)* | -408723<br>(P=0.1419) | 250991<br>(P<0.001)*** | 2954162 | 222% |
| Appointments Booked<sup>1</sup> | -6196 *<br>(P=0.02) | 2247***<br>(P<0.001) | -21315***<br>(P<0.001) | -22<br>(P=0.95) | -21601 | -38% |
| Appointments Booked<sup>2</sup> | -6196*<br>(P=0.02) | 2247***<br>(P<0.001) | -20294***<br>(P<0.001) | -226<br>(P=0.62) | -23236 | -41% |

p-values: 0 '\*\*\*' 0.001 '\*\*' 0.01 '\*' 0.05 '.' 0.1 ' ' 1
<sup>1</sup> January 2019-May 2021
<sup>2</sup> January 2019-April 2021

**Table 3.** GP Health Records Viewed & Prescriptions Ordered Interrupted Times Series Output.

| NHS App Metric | Regression Intercept<br>(p-value) | Pre-Intervention Trend<br>(p-value) | Immediate Change in Trend after COVID-19 Lockdown<br>(p-value) | Change in Trend over Study Period<br>(p-value) | 1 year after first lockdown |  |
| --- | --- | --- | --- | --- | --- | --- |
|  |  |  |  |  | Absolute Change<br>(count) | Relative Change<br>(percent) |
| GP Health Records Viewed<sup>1</sup> | -86104<br>(P=0.89) | 25159<br>(P=0.72251) | -1441297<br>(P=0.11) | 371656**<br>(P=0.001) | 3390234 | 548% |
| GP Health Records Viewed<sup>2</sup> | -86104<br>(P=0.56) | 25159<br>(P=0.1349) | -495304*<br>(P=0.02) | 182458<br>(P<0.001)*** | 1876645 | 303% |
| Prescriptions Ordered<sup>1</sup> | -18935<br>(P=0.17) | 5225 **<br>(P=0.001) | 27494<br>(P=0.15) | 19934 ***<br>(P<0.001) | 286630 | 225% |
| Prescriptions Ordered<sup>2</sup> | -18935*<br>(P=0.01) | 5225 ***<br>(P<0.001) | 45036<br>(P<0.001) | 16425<br>(P<0.001) | 258563 | 202% |

**p-values: 0 '\*\*\*' 0.001 '\*\*' 0.01 '\*' 0.05 '.' 0.1 ' ' 1**
<sup>1</sup> January 2019-May 2021
<sup>2</sup> January 2019-April 2021

#### Prescriptions Ordered

Repeat prescriptions ordered using the NHS App ranged from 655 to 530,382 per month from January 2019 to May 2021. There was a 1.3 times increase in prescriptions ordered after announcement of the first lockdown, March 2020 (98,692 prescriptions ordered) and April 2020 (128,028 prescriptions ordered) (**Figure 4**). The ITS showed that pre-lockdown, there was significant evidence that the number of prescriptions ordered was increasing at an average rate of 5,255 (p=0.001) a month. (**Table 2**). Post-lockdown, there was a positive and significant increase in trend of 19,934 (p<0.001) prescription orders per month. (**Table 2**). 12 months after the first lockdown, the average number of NHS App prescription orders was 286,630 more than would have been expected if lockdown did not occur. This represented a 225% increase.

## Discussion

### Principal Findings

Our observational analysis found that there has been strong adoption of the NHS App even before the onset of the COVID-19 pandemic. Prior to when the first national lockdown in the UK was announced, there were almost 1.5 million downloads of the App. From April 2021 to May 2021, there was a 4-fold increase in App downloads, showing the significant impact the introduction of the COVID Pass had on App uptake. However, a disparity between App downloads and registrations still exists. At the end of the study period, in May 2021, there were 8,524,882 downloads and 4,446,286 registrations. This could be for several reasons, from users not being able to successfully register for the App, or never using the App once it’s downloaded. A recent study on mobile app abandonment rate found that 25% of all mobile apps were only accessed once after download.^**16**^

COVID-19 and the introduction of the COVID Pass service had significant impacts on different functionalities of the App. Introduction of the COVID Pass accounted for over a 3 fold increase in login sessions in May, indicating that users could have been logging into the App to retrieve their vaccination status. This pattern was also observed for the number of users using the App to access their GP health records. Appointment bookings fell substantially after the first lockdown but continued on the same gradient from March-May 2021. These findings support current events at the time where in person GP appointments came to a standstill due to the pandemic, and some GPs switched off the ability for patients to book appointments via the NHS App at this time. Prescriptions ordered via the NHS App also significantly increased after the first lockdown, suggesting that more users were utilizing the App to place prescription orders rather than going in person or via the phone. These findings could be related to patients and carers being less likely to seek healthcare during the pandemic, in support of the pressure experienced by NHS services at the time, not wanting to bother the health service, and seeking alternative means to manage their health, either through self-management, or remote access (e.g. digital prescription ordering), as well as fear of getting infected.^**27**^

App registrations across England varied by CCG, and no regional patterns were distinctly observed. Regions with the lowest proportion of App registrations seemed to be in some of the more deprived parts of England, which would merit further exploration at the level of individual data.

The public health impact of digital health interventions is dependent upon real-world uptake, and engagement.^**17**^ Our analysis suggests that there is high uptake of the NHS App, but this may not always correspond with high levels of engagement. A recent study looking at uptake and engagement of health and well-being apps found that one of most important factors for engagement were apps coupled with health practitioner support.^**17**^ Apps with a human support component were found to be more effective in increasing the effectiveness and engagement of digital health interventions.^**17**^ At the time of this analysis, the NHS App does not directly incorporate any health practitioner support.

#### Remaining Gaps and Limitations

The results of this observational study indicate that more research is needed to identify whether the NHS App is an effective digital health tool and the extent to which it has met the goals set out by NHS England. The existing literature on technology adoption and diffusion of innovation tells us that this process is difficult and complex. Adoption is not just based on the technology, but a complex mixture of how the public and staff interact with it, what they see as the benefits, organizational culture and wider influences on the system including the policy and regulatory context.^**25**^ Our analysis showed that there has been strong adoption of the App, and that COVID has significantly expedited uptake, however, further research is needed to evaluate continuous usage of the App and if it yields any benefits.

Our ecological analysis was only able to analyze the NHS App at the GP practice level, which did not allow us to make any inferences on individual app usage behavior. Having data only at the GP practice level data did not allow us to explore whether different subgroups were impacted by the App differently. Previous studies have shown that ethnic minorities, patients who are younger, healthier, and less educated were less likely to adopt to patient portals.^14^ Person-level data of NHS App usage is needed to see if these same results are observed. Having person-level data would also further our evaluation by allowing us to assess whether there are any demographic and socio-demographic variations within App usage, and if the intervention has had any impact on clinical outcomes. The interrupted time series analysis also does not account for other confounding factors that may have contributed to the results seen in this study. This could include changes in seasonality, other existing co-interventions during the study period, and other events that may have occurred during the intervention. We tried to minimize known confounders by analyzing the data with and without the introduction of the COVID Pass, which has been included in the appendix. We intend to look at long term App usage once more data is available post-COVID.

As part of this NIHR-funded study, we are also conducting a qualitative assessment to understand the multiple interacting influences on uptake and use of the app from the perspective of patients, healthcare staff and commissioning and policy stakeholders.

## Conclusion

This is the first ecological study that has analyzed a nationwide intervention rolled out by NHS England. This analysis has shown that uptake and adoption of the NHS App is high, and has been driven by COVID related events, but further research is needed to measure the extent to which it improves patient experience and influences health service access and care outcomes. These results may inform other initiatives to harness patient-facing digital tools, and suggest improvements on future digital health interventions.

## Data Availability

All data produced in the present study are available upon reasonable request to the authors

https://digital.nhs.uk/services/nhs-app/nhs-app-dashboard

