## Appendix 1 for "An observational study of uptake and adoption of the NHS App in England"

**Table 1: Summary of NHS app data metrics available from the NHS App Dashboard**

| Metric Name | Granularity | Description |
|---|---|---|
| NHS App Registrations | GP practice | Number of App users who have obtained P9 verification (full access) in the specified reporting period |
| NHS App downloads | National | Number of NHS App downloads from Google and Apple Stores in the specified reporting period |
| Total GP registered patients aged 13+ years | GP practice | Number of patients registered at a general practice to date (cumulative) |
| NHS App Logins | GP practice | Number of NHS App logins in the specified reporting period |
| Users Booking Appointments | GP practice | Number of GP appointments booked in the specified reporting period |
| Appointment Cancellation | GP practice | Number of GP appointments canceled in the specified reporting period |
| Users Requesting Prescriptions | GP practice | Number of prescriptions requested in the specified reporting period |
| User Accessing GP health records | GP practice | Number of GP health records accessed in the specified reporting period |
| Users Visiting NHS 111 Online | National | Number of App users visiting NHS 111 online from the NHS App in the specified reporting period |
| Organ Donation Registrations | GP practice | Number of organ donor registrations in the specified reporting period |
| Organ Donation Updates | GP practice | Number of organ donation status updates in the specified reporting period |
| Organ Donation Withdrawals | GP practice | Number of organ donor registration withdrawals in the specified reporting period |
| Users Visiting Health A-Z | National | Number of App users visiting the Health A-Z page in the specified reporting period |

| | | |
|---|---|---|
| Users visiting National Data Opt-out | National | Number of App users visiting the National Data opt-out page in the specified reporting period |

### Graph 1: Login Sessions & Appointments Booked Interrupted Times Series-Without May 2021 data

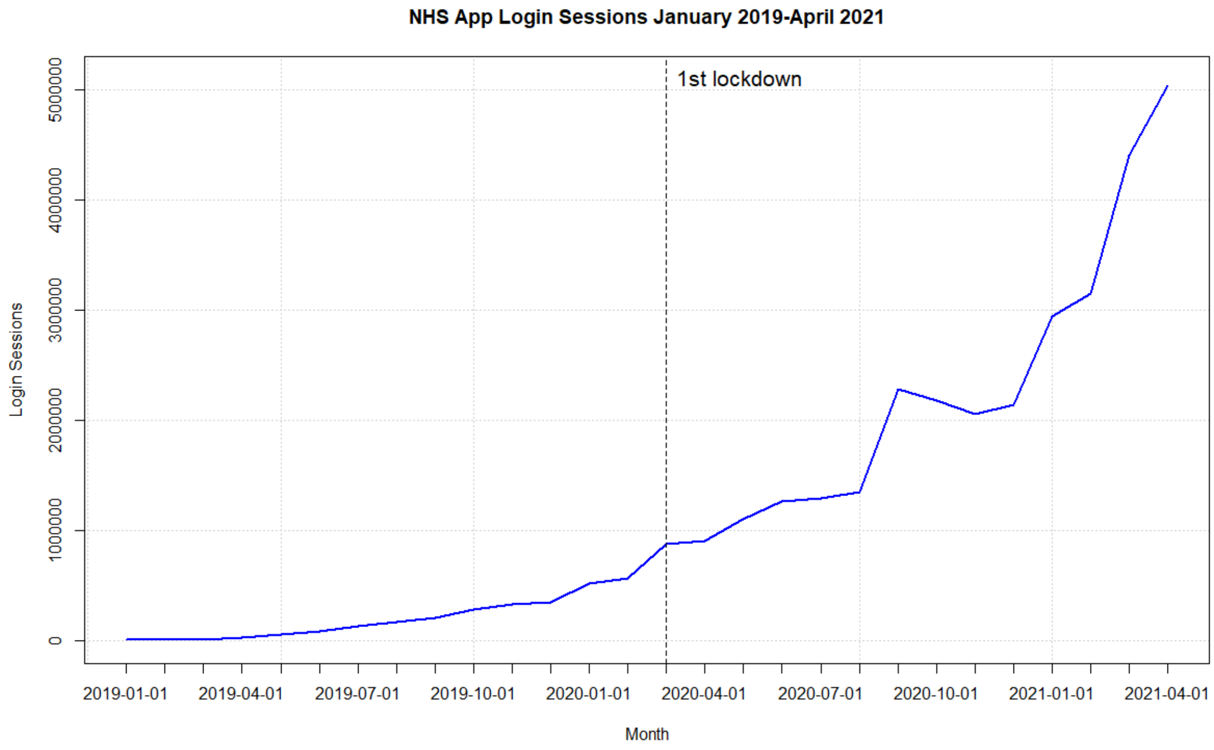


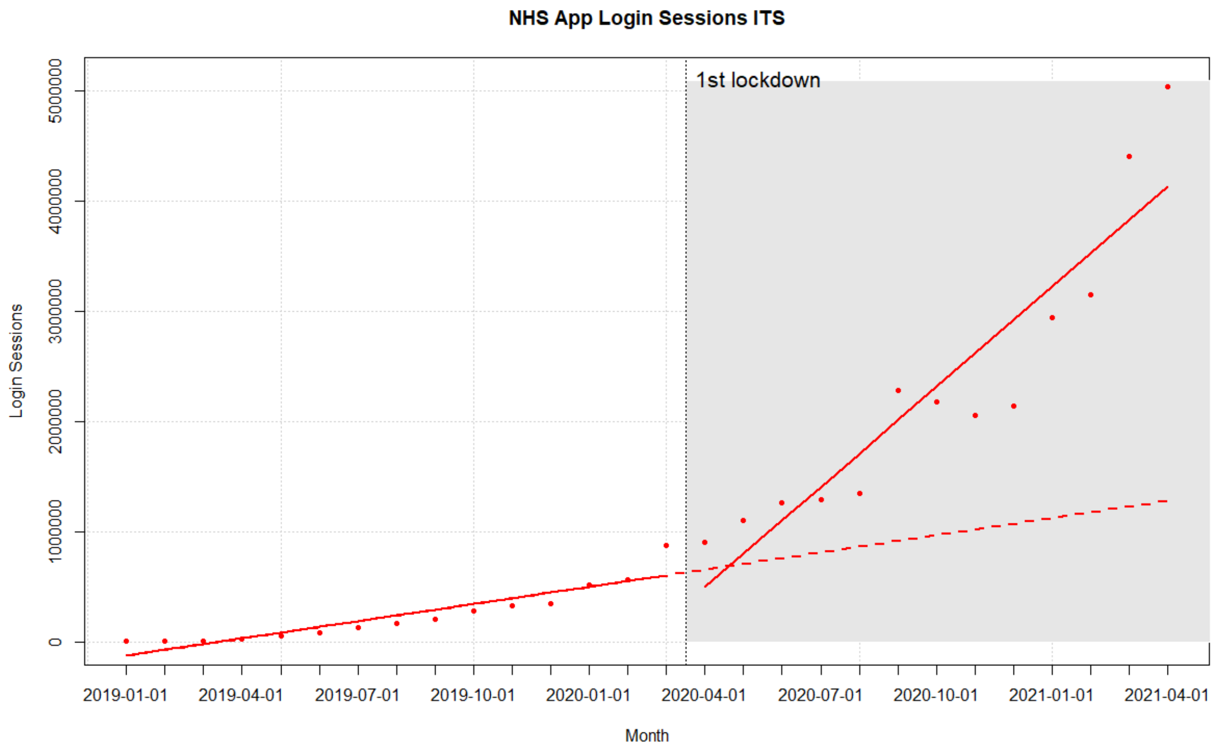

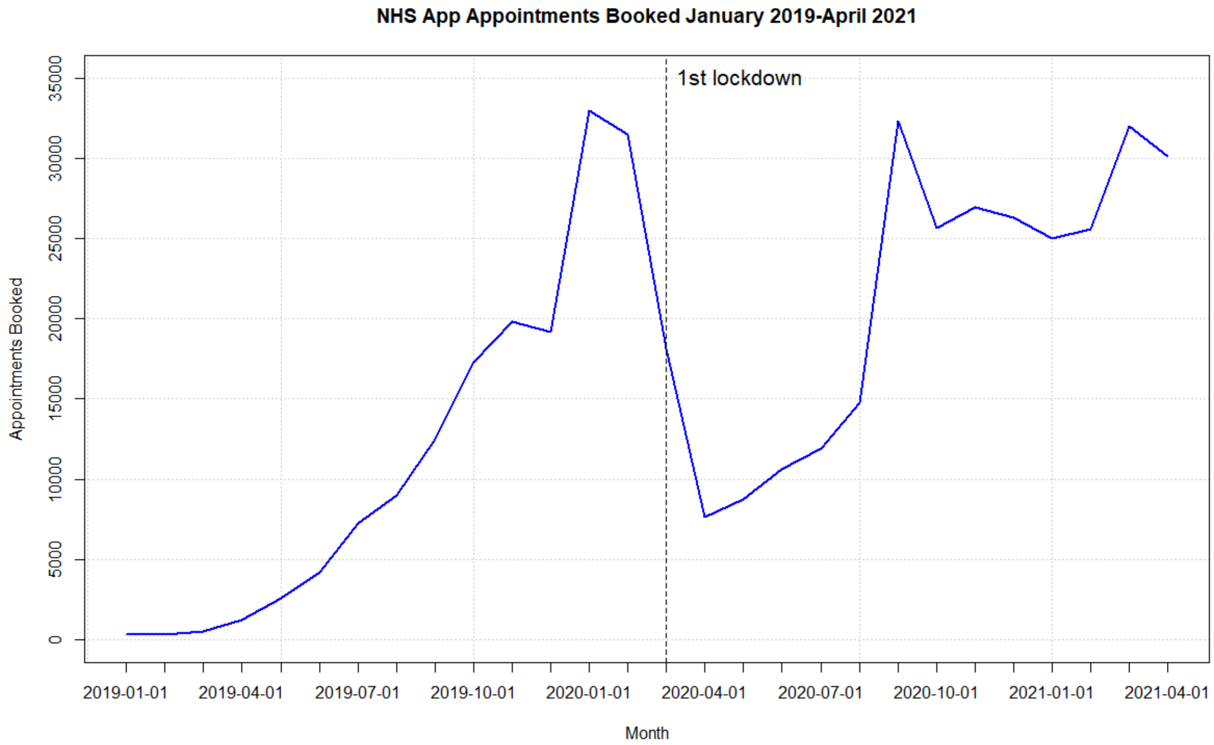
NHS App Appointments Booked January 2019-April 2021
1st lockdown
Appointments Booked
0
5000
10000
15000
20000
25000
30000
35000
2019-01-01
2019-04-01
2019-07-01
2019-10-01
2020-01-01
2020-04-01
2020-07-01
2020-10-01
2021-01-01
2021-04-01
Month

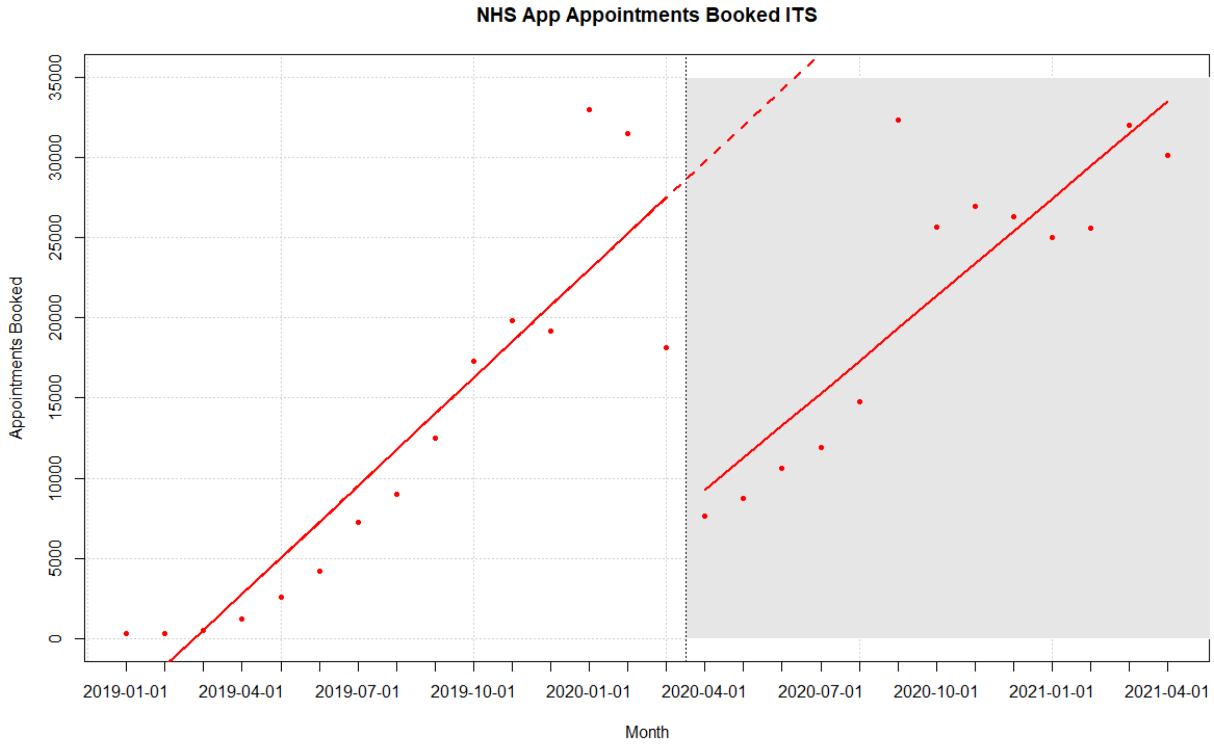
NHS App Appointments Booked ITS
Appointments Booked
0
5000
10000
15000
20000
25000
30000
35000
2019-01-01
2019-04-01
2019-07-01
2019-10-01
2020-01-01
2020-04-01
2020-07-01
2020-10-01
2021-01-01
2021-04-01
Month

**Graph 2: GP Health Records Views & Prescriptions Ordered Interrupted Times Series-Without May 2021 data**

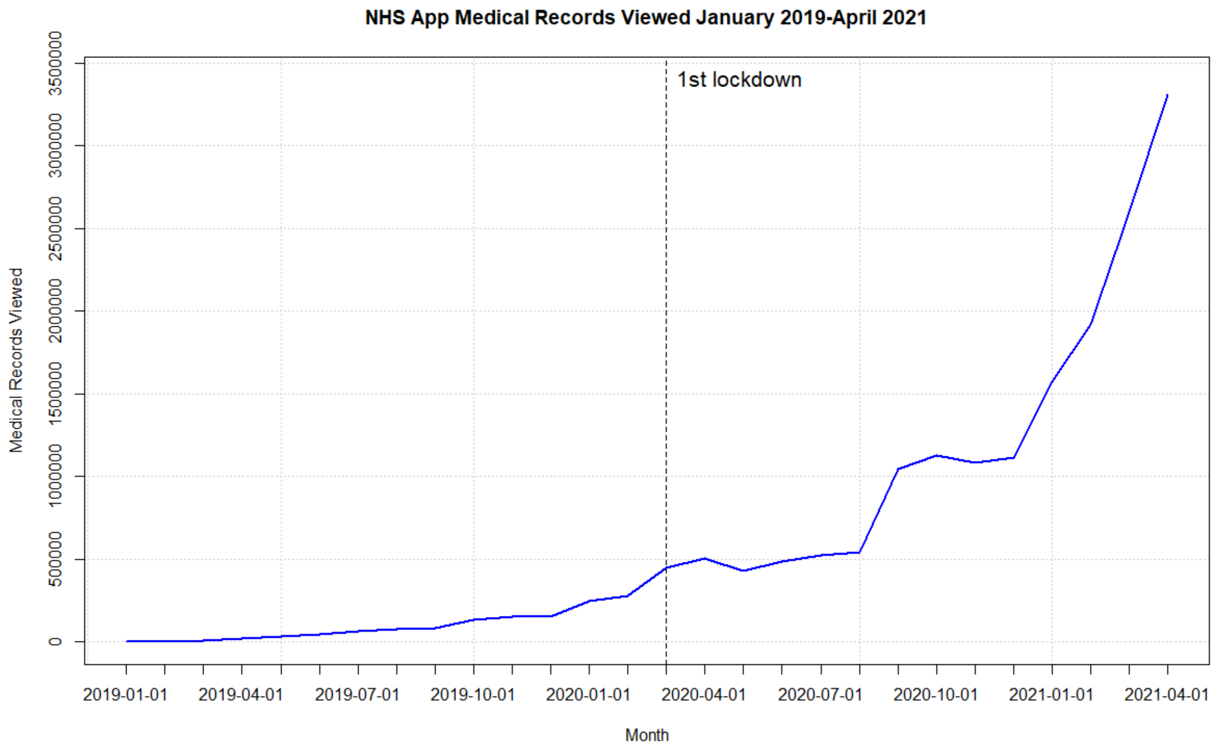


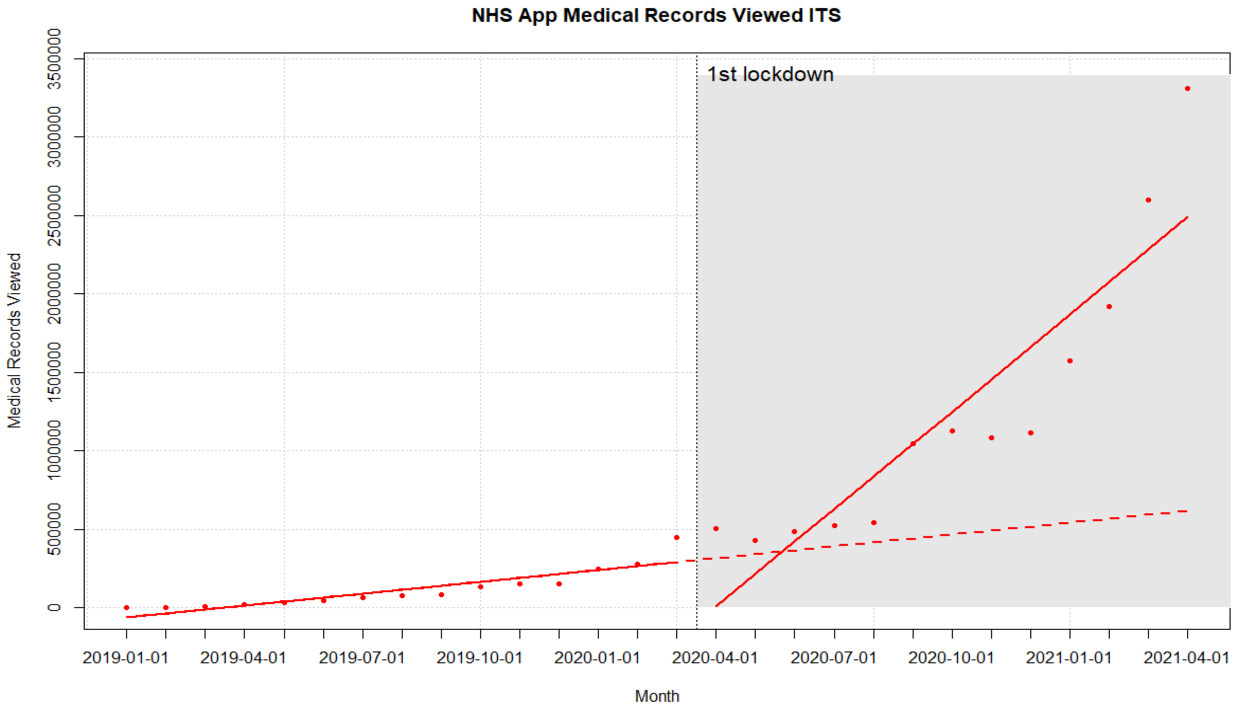

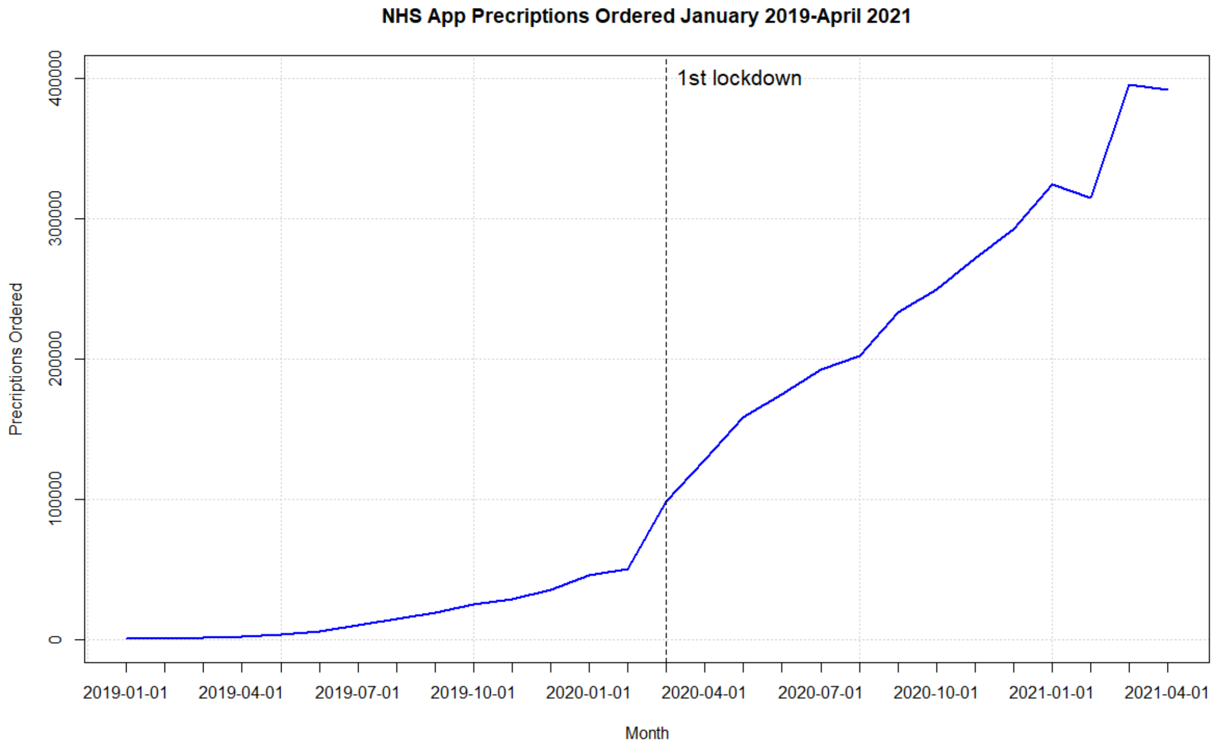

NHS App Precriptions Ordered January 2019-April 2021
1st lockdown
Precriptions Ordered
0
100000
200000
300000
400000
2019-01-01
2019-04-01
2019-07-01
2019-10-01
2020-01-01
2020-04-01
2020-07-01
2020-10-01
2021-01-01
2021-04-01
Month


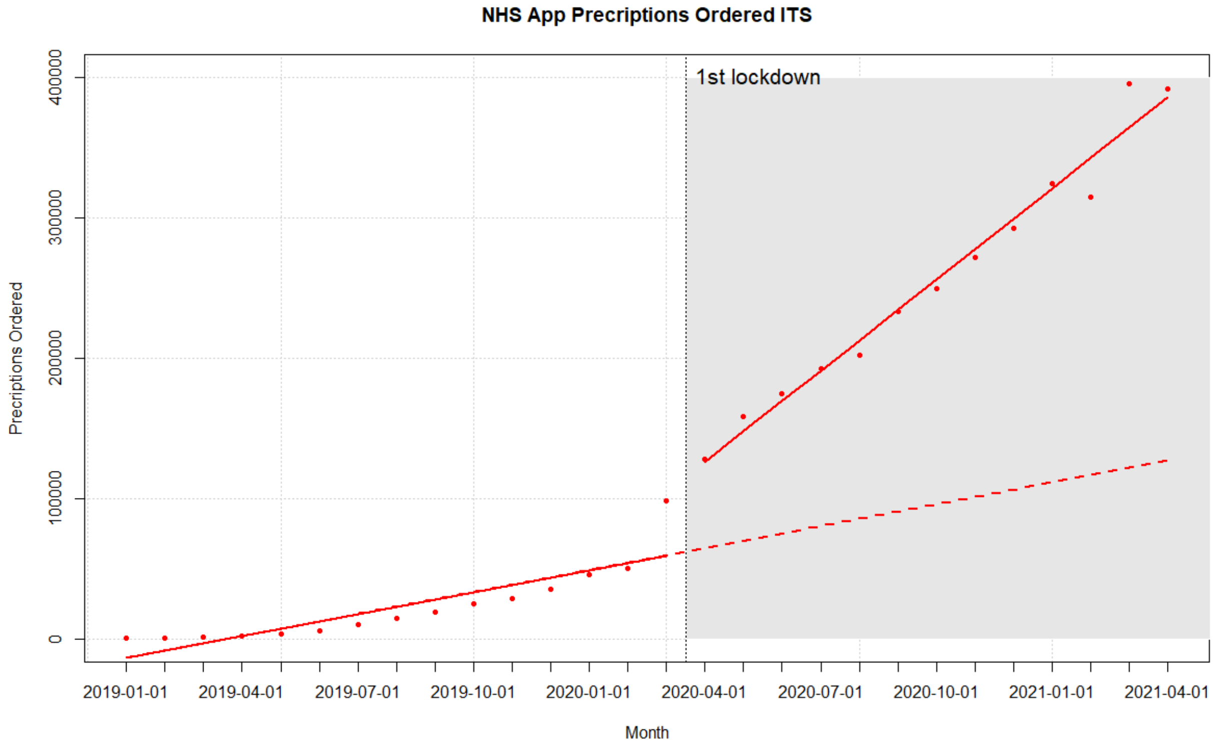

NHS App Precriptions Ordered ITS
1st lockdown
Precriptions Ordered
0
100000
200000
300000
400000
2019-01-01
2019-04-01
2019-07-01
2019-10-01
2020-01-01
2020-04-01
2020-07-01
2020-10-01
2021-01-01
2021-04-01
Month
